# Management of mild COVID-19: Policy implications of initial experience in India

**DOI:** 10.1101/2020.05.20.20107664

**Authors:** Rohit Kumar, Bisakh Bhattacharya, Ved Prakash Meena, Anivita Aggarwal, Manasi Tripathi, Manish Soneja, Ankit Mittal, Komal Singh, Nishkarsh Gupta, Rakesh Kumar Garg, Brajesh Kumar Ratre, Balbir Kumar, Shweta Arun Bhopale, Pavan Tiwari, Ankit Verma, Sushma Bhatnagar, Anant Mohan, Naveet Wig, Randeep Guleria

**Affiliations:** Department of Medicine, All India Institute of Medical Sciences, New Delhi (110029); Department of Ophthalmology, All India Institute of Medical Sciences, New Delhi (110029); Department of Onco-anesthesia & Palliative Medicine, All India Institute of Medical Sciences, New Delhi (110029); Department of Pulmonary Medicine & Sleep Disorders, All India Institute of Medical Sciences, New Delhi (110029); Department of Pediatrics, All India Institute of Medical Sciences, New Delhi (110029)

**Author notes:** **corresponding author** Dr. Manish Soneja, Additional professor, Department of Medicine, AIIMS, Medicine office, 3rd floor, Teaching block, AIIMS, New Delhi (110029), Email id.

## Abstract

**Objectives:** Ongoing pandemic due to COVID-19 has spread across countries, surprisingly with variable clinical characteristics and outcomes. This study was aimed at describing clinical characteristics and outcomes of admitted patients with mild COVID-19 illness in the initial phase of pandemic in India.

**Design:** Retrospective (observational) study.

**Setting:** COVID facilities under AIIMS, New Delhi, where, isolation facilities were designed to manage patients with mild illness and dedicated COVID ICUs was created to cater patients with moderate to severe illness.

**Participants:** Patients aged 18 years or more, with confirmed illness were eligible for enrolment. Patients who were either asymptomatic or mildly ill at presentation were included. Patients with moderate to severe illness at admission, or incomplete clinical symptomatology records were excluded.

**Methods:** Data regarding demographic profile, comorbidities, clinical features, hospital course, treatment, details of results of RT-PCR for SARS-CoV-2 done at baseline and at day 14, chest radiographs (wherever available) as well as laboratory parameters was obtained retrospectively from the hospital records.

**Main outcome measures:** Final outcome was noted in terms of course of the disease, patients discharged, still admitted (at time of conclusion of study) or death.

**Results:** Out of 231 cases included, majority were males(78·3%) with a mean age of 39·8 years. Comorbidities were present in 21·2% of patients, diabetes mellitus and hypertension being most common. The most common symptoms were dry cough(81, 35%), fever(64, 27·7%), sore throat(36, 15·6%), and dyspnoea(24, 10·4%); asymptomatic infection was noted in 108(46.8%) patients. Presence of comorbidities was an independent predictor of symptomatic disease (OR- 2·66; 95% CI 1·08 to 6·53, p= 0·03). None of the patients progressed to moderate to severe COVID-19. There were no deaths in this cohort.

**Conclusions:** Patients with mild disease at presentation had a stable disease course and therefore such cases can be managed outside hospital setting. A large proportion of patients remained asymptomatic throughout the course of infection and those with comorbidities are more likely to be symptomatic.

**Trial registration:** Not applicable

## Introduction

The SARS-CoV-2 emerged towards the end of 2019 and since then it has spread across the globe. The coronavirus disease 2019 (COVID-19) pandemic has been raging across different geographies and a wide range of morbidity and mortality rates has been reported from various settings with case fatality rates ranging from 1·8-15%. ^(1–3)^ This variation can be explained by the age and comorbidity profile of the affected population, testing criteria and admission criteria, besides other factors. ^(4-6)^ To have a complete understanding of the spectrum of illness, severity of disease and outcomes; studies are required which includes confirmed patients with mild illness and asymptomatic close contacts of positive cases, besides the moderate to severe cases which require hospitalization. This will be useful for policymakers to decide on actions required for control of the pandemic and optimum utilization of healthcare resources in resource-limited settings in particular.

First case of COVID-19 from India was reported on January 30, 2020 from Kerala. The cases in India began rising in the later half of the month of March. The Indian authorities enforced lockdown quite early in the course of the pandemic i.e. on March, 25. Large scale testing and contact tracing was carried out. All patients were admitted irrespective of symptomatology and their close contacts were admitted in isolation facilities and tested for SARS-CoV-2. Subsequently, the guidelines were modified to allow home isolation of very mild/ pre-symptomatics COVID-19 cases. This initial phase presented an unique opportunity to elucidate the disease profile and outcomes among patients presenting with mild symptoms and those diagnosed on contact tracing.

This study was designed to study the clinical characteristics and outcomes including progression to moderate-severe disease among patients with mild COVID-19.

## Methods

### Study design and setting

This was a retrospective study conducted in COVID-19 facilities at AIIMS, New Delhi among patients admitted between 20th March and 30th April 2020. It is a tertiary care teaching hospital wherein, facilities for management of COVID-19 patients were created. Isolation facilities were designed to manage patients with mild illness and a dedicated COVID ICU was created to cater patients with moderate to severe illness. Patients presenting to the facility included those with varying degrees of disease severity as well as asymptomatic close contacts of a large congregation, wherein a number of positive cases were reported.

### Procedures

The clinical case records of all admitted patients with laboratory-confirmed COVID-19 were screened for eligibility. The diagnosis was confirmed by real-time reverse-transcriptase-polymerase-chain-reaction (RT-PCR) for detection of SARS CoV-2 on the specimens obtained from nasal and throat swabs. Patients of age more than 18 years and asymptomatic/ mild illness at presentation were included. Patients with incomplete records of clinical symptomatology were excluded. Patients were classified as having mild disease if they presented with uncomplicated upper respiratory tract viral infection with non-specific symptoms such as fever, fatigue, cough, etc or features of pneumonia but no signs of severe pneumonia and no need for supplemental oxygen.^(7)^

Data regarding demographic profile, comorbidities, clinical features, laboratory parameters, hospital course and treatment outcomes were noted. The daily symptom screen included fever, cough, sore throat, shortness of breath, expectoration, rhinorrhoea, myalgia, fatigue, loose stools, chest pain, anorexia, loss of taste and loss of smell and other symptoms as given by the patients. Patients who did not have any symptoms related to COVID-19 at admission or during the hospital course were considered asymptomatic. Details of results of RT-PCR for SARS-CoV-2 done at baseline and at day 14 was obtained retrospectively from the hospital records. Chest radiographs (wherever available) as well as available laboratory parameters were assessed for any abnormality.

Patients were managed as per the national guidelines.^(8)^ In case of clinical deterioration, the protocol was to shift patients to a high dependency unit (HDU)/ intensive care unit (ICU) for further management. Patients were considered dischargeable after 2 negative SARS-COV-2 RT PCR reports (at least 24 hours apart), performed after at least 7 days of last positive test results. Final outcome was noted in terms of number of patients discharged, patients still admitted (at time of conclusion of study) or death. The permission for the study was obtained from the Institute Ethics Committee.

### Statistical analysis

Data was analysed using STATA 13·0. The data is presented as mean (standard deviation)/ median (interquartile range) and frequency percentage. Normality of data was assessed by Shapiro-Wilk’s test. Logistic regression analysis was used to find out independent predictors for symptomatic illness and RT-PCR negativity at 14 days. p-value < 0·05 was considered as statistically significant.

## Results

The study was conducted in the initial part of the pandemic when patients irrespective of severity of symptomatology were admitted. A total of 335 laboratory-confirmed COVID-19 patients were admitted to the facility in the study period. Among them, 231 cases were included after applying the inclusion and exclusion criteria (Fig. 1).

**Figure 1.**
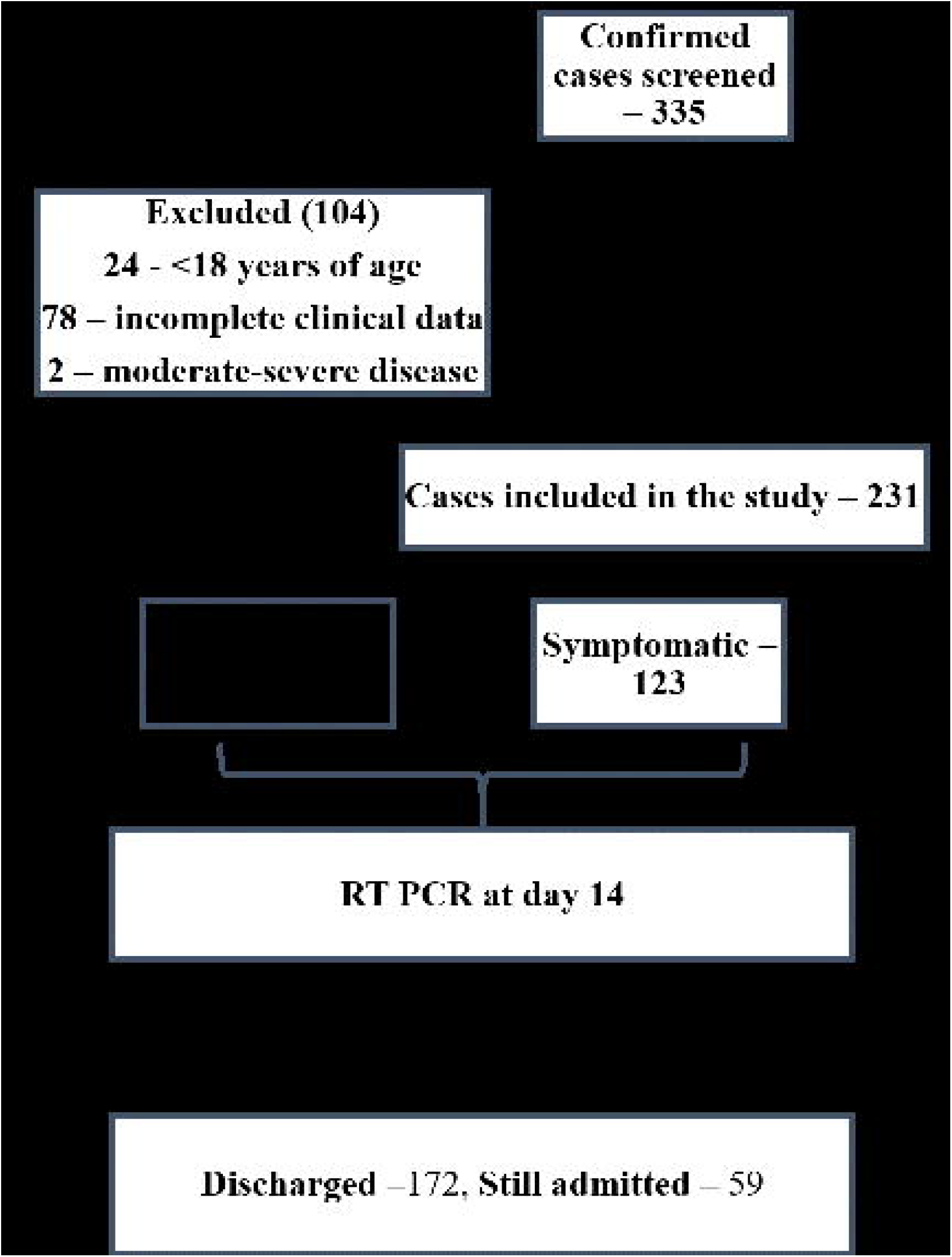
Study flow diagram

Majority were males comprising 78·4% of the cohort with mean age 39·8 years (range 18-73 years) (Table 1). Most patients (205, 88·7%) had a definitive history of contact with a suspected or confirmed case. A significant proportion of the patients (49, 21·2%) had one or more chronic diseases including diabetes mellitus, hypertension, cardiovascular or cerebrovascular disease, chronic lung disease including asthma/ chronic obstructive pulmonary disease /tuberculosis or chronic liver disease. Diabetes mellitus (28, 12.1%) and hypertension (19, 8.2%) were the most common comorbidities noted.

**Table 1:**
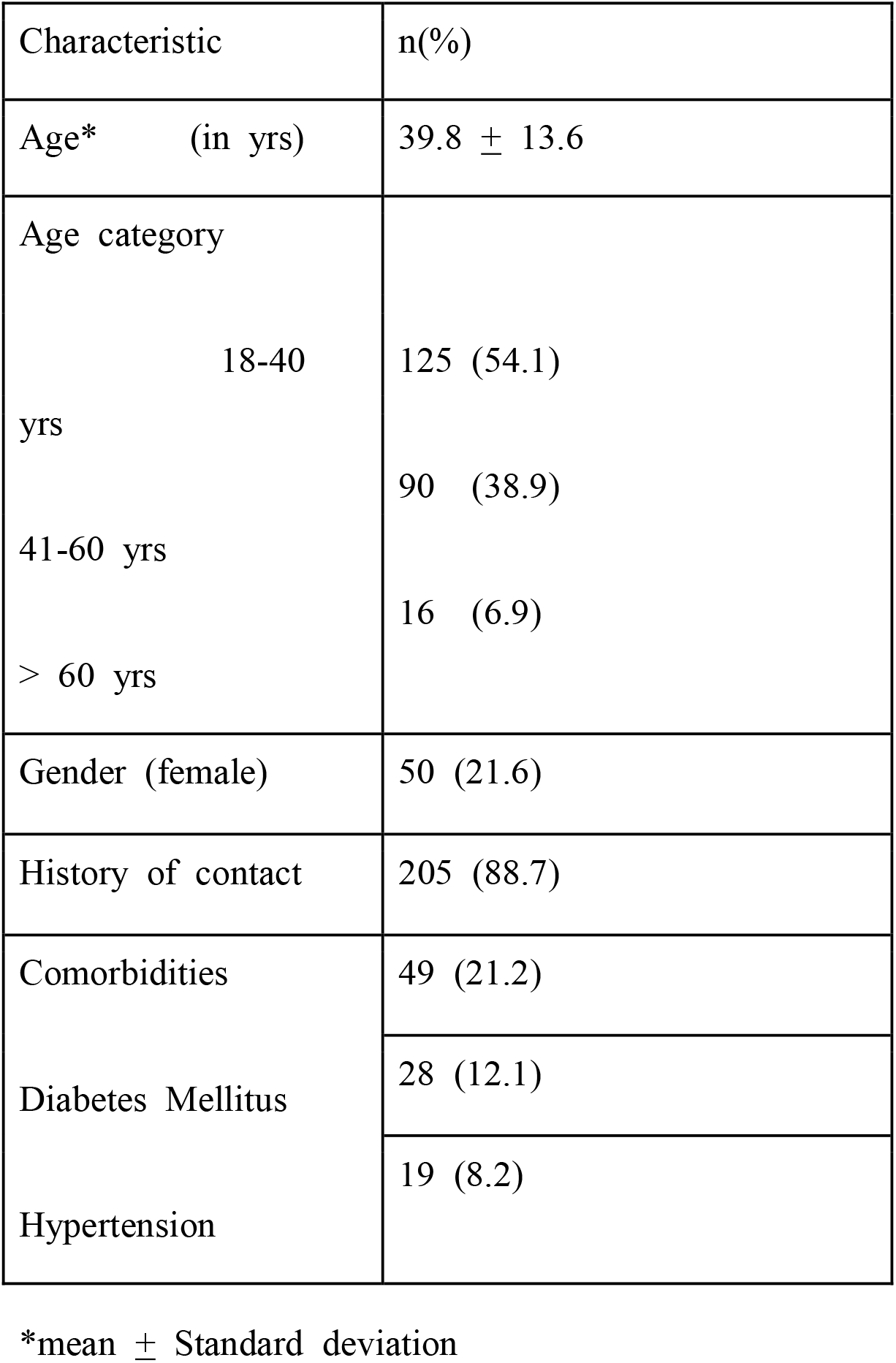
Baseline characteristics of COVID-19 patients (n=231)

Symptoms specific to COVID-19 were present in 123 (53·2%) of the patients; whereas, 108 (46·7%) patients remained asymptomatic. The median duration of symptoms was 3 days (IQR 2-5·5). Among symptomatics, the most common symptoms included dry cough (81, 65·8%), fever (64, 52%), sore throat (36, 29·2%) and shortness of breath (24, 19·5%)(Figure 2). The most common presentation among symptomatic patients was with dry cough alone (27, 11·7%). Other common presenting features were fever, fever with dry cough; and fever with cough and shortness of breath (Figure 3). The laboratory abnormalities included anemia in 12·2%, leukopenia in 4·8% and lymphocytopenia (<1500 cells/cumm) in 11·2% of the patients.

**Figure 2.**
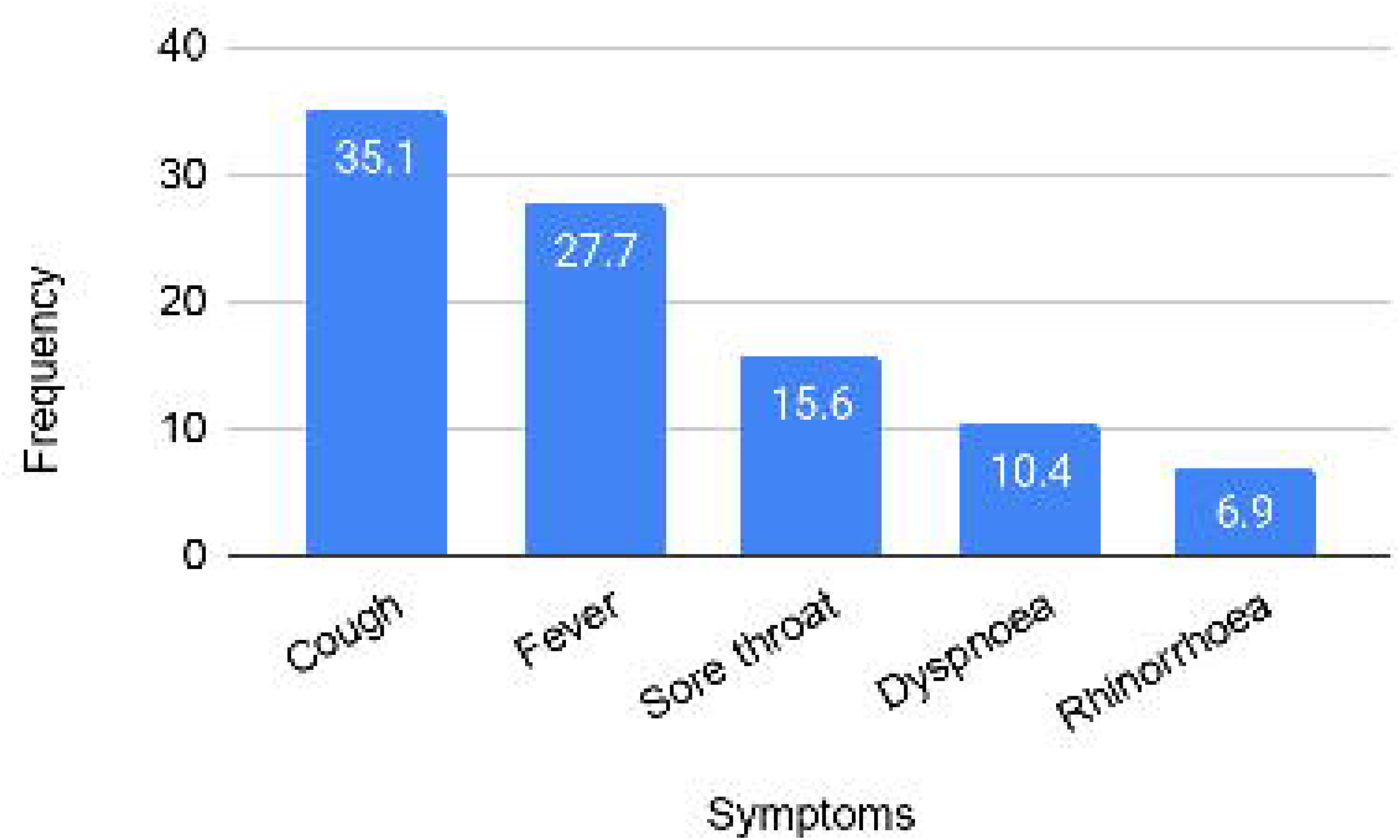
Frequency of common symptoms among COVID-19 patients (n=231)

**Figure 3.**
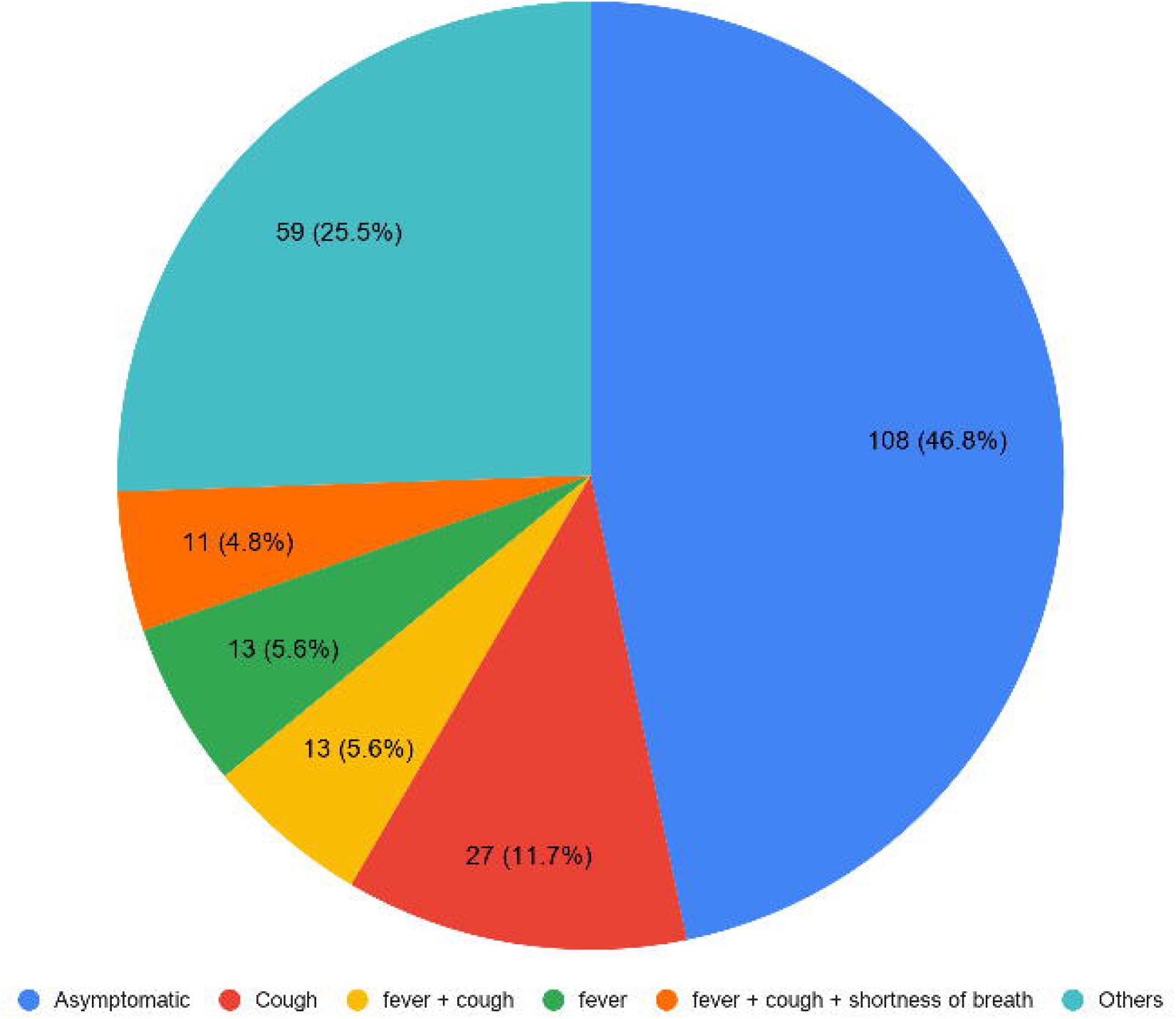
Symptomatology among COVID-19 patients (n=231)

The demographic and laboratory parameters are compared between symptomatic and asymptomatic patients in table 2. Presence of comorbidities emerged as an independent predictor of symptomatic disease with odds ratio of 2·66 (95% CI 1·08-6·53, p= 0·03) on logistic regression analysis.

**Table 2:**
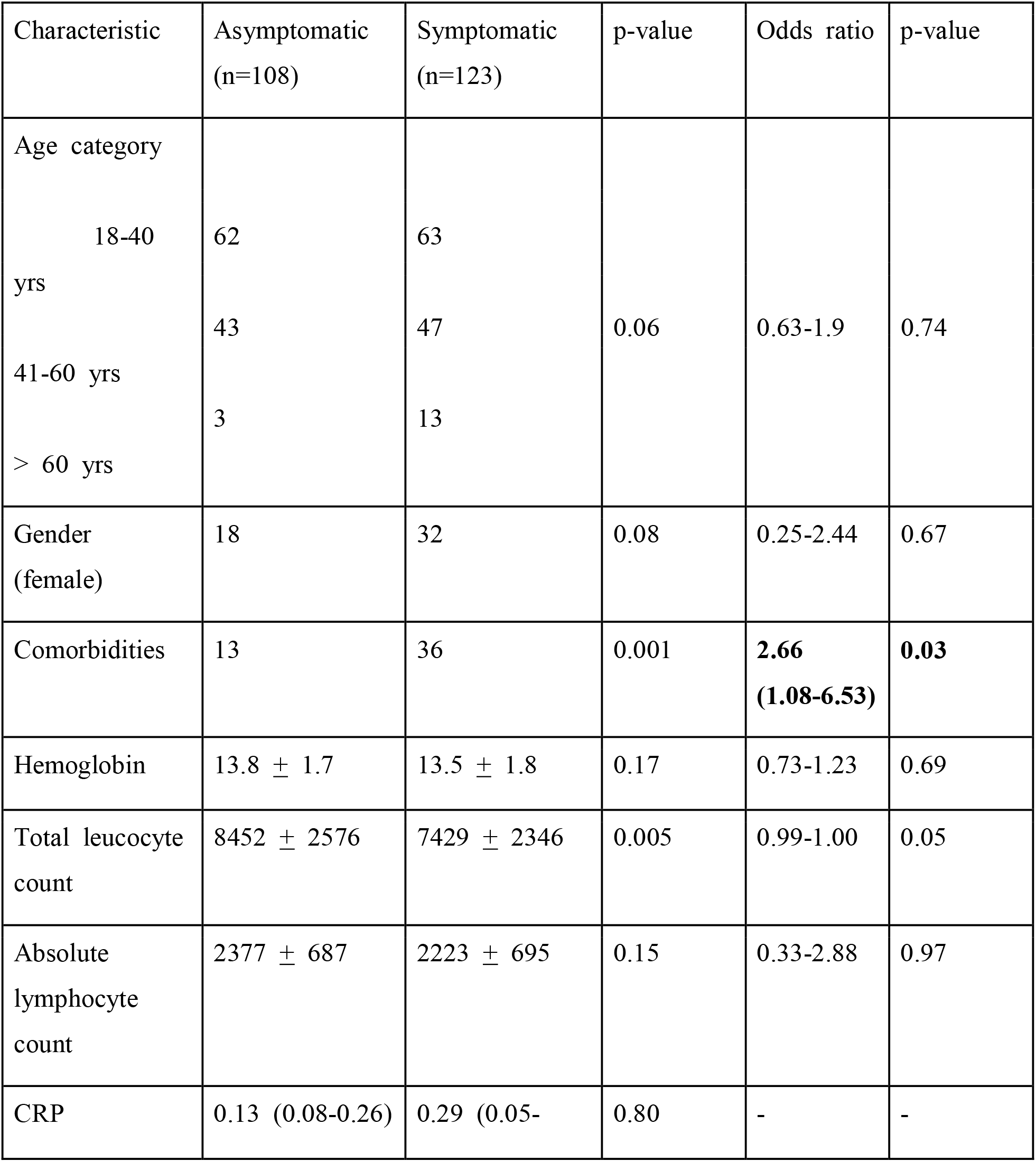
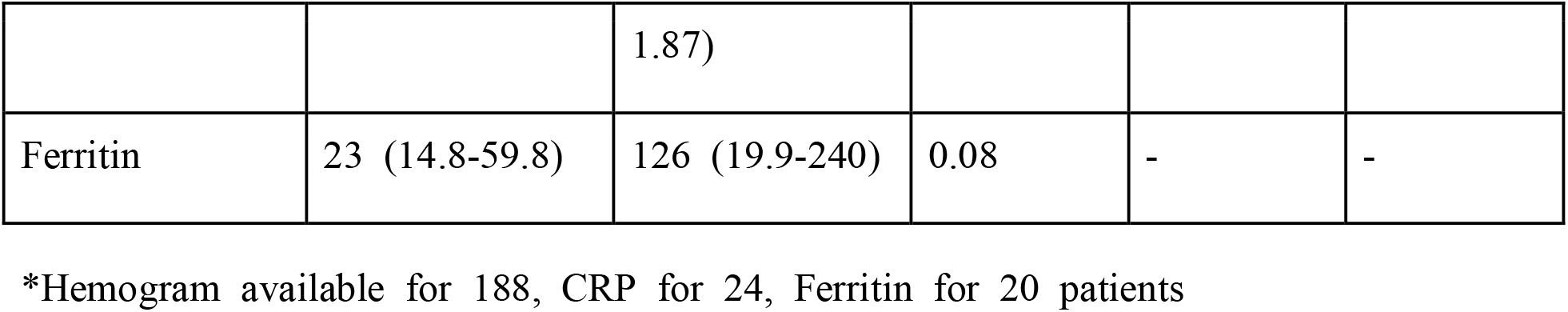
Demographic and laboratory parameters among symptomatic and asymptomatic patients (n=231)

RT-PCR for SARS-CoV-2 was positive in 14·5% (26/179) of patients at the end of 2 weeks. Table 3 shows the comparison of various demographic, clinical and laboratory parameters among these 2 groups of patients, wherein no statistically significant differences were noted.

**Table 3:** Demographic and laboratory parameters among RT-PCR positive and negative patients at the end of 2 weeks (n=179)

| Characteristic | RT-PCR positive<br>(n=26) | RT-PCR<br>negative(n=153) | p-value | Odds<br>ratio | p-value |
| --- | --- | --- | --- | --- | --- |
| Age category |  |  |  |  |  |
| 18-40 yrs | 13 | 81 | 0.77 | 0.52-2.31 | 0.80 |
| 41-60 yrs | 11 | 61 |  |  |  |
| > 60 yrs | 2 | 11 |  |  |  |
| Gender<br>(female) | 6 | 19 | 0.14 | 0.10-1.61 | 0.20 |
| Comorbidities | 5 | 30 | 0.96 | 0.20-2.78 | 0.67 |
| Symptomatic | 12 | 72 | 0.93 | 0.43-2.55 | 0.9 |
| Received HCQ | 6 | 29 | 0.62 | 0.41-4.02 | 0.65 |
| Hemoglobin | 13.6 ± 1.9 | 13.8 ± 1.5 | 0.59 | 0.77-1.46 | 0.7 |
| Lymphopenia<br>(<1500/mm<sup>3</sup>) | 2 | 12 | 0.97 | 0.17-6.22 | 0.97 |

Among the 231 cases none progressed to moderate-severe disease during the course of hospitalization. Four patients (1.7%) required shifting to the ward unit to manage comorbidities including angina (1), ureteric colic (1) and glycemic control (2). Overall, 8-1% (4/49) patients with comorbidities required uptriaging. There were no deaths in this cohort.

At the time of writing this manuscript, 172 (74·5%) patients have been discharged and 59 (25·5%) are still admitted.

## Discussion

In this retrospective study, 231 admitted patients with mild illness were recruited. The mean age was 40 years and one-fifth had some sort of comorbidities. Cough (65·8%) and fever (52%) were the most common features among symptomatics. A large proportion of patients (46·7%) remained asymptomatic throughout the course of the infection; patients with comorbidities were more likely to be symptomatic. None of the patients progressed to moderate-severe disease.

The cohort we recruited is younger compared to most studies;^(5,9-12)^ but similar in age profile of few reported studies.^(13,14)^ This is an important context to interpret the results because increasing age and associated comorbidities are well known risk factors for disease severity and outcomes.^(5,10,15-17)^ About one-fifth of our patients had comorbidities which is higher than reported among mild cases in the same age-group from Europe; ^(13)^ however, it is closer to the reports among higher age-groups from other regions.^(9)^ This probably reflects the increased burden of noncommunicable diseases in Indian population.^(18)^ Thus our cohort was unique in being younger but with a higher burden of comorbidities compared to other reports of COVID-19.

The reports on symptomatology are varied across geographies. Fever and cough are the most common symptoms reported across various countries.^(9,15,19,20)^ In our study cough and fever were the commonest symptoms, with former being more common. It has important implications because patients with mild cough and no fever may ignore their symptoms and not seek medical attention. Further, the frequency of these symptoms were much lower than reported in other studies. In a study from China, the reported frequencies of fever and cough was reported as 88% and 68% respectively, whereas in the United States it was 94% and 88%.^(9,20)^ It is in stark contrast to 35% and 28% seen in the present study. It may be related to the diagnostic strategy and admission criteria which is variable across countries. This study was carried out in early days of the pandemic in India when large scale surveillance, testing and contact tracing was carried out. All patients were admitted irrespective of symptomatology and their close contacts were admitted in isolation facilities and tested for SARS-CoV-2. This presented a unique opportunity to study the disease course in all infected patients irrespective of symptomatology.

A significant proportion of our patients were asymptomatic throughout the course of admission. The literature on asymptomatic infections are sparse at present. This is likely due to the fact that these patients would not seek medical care or may have been treated at home as the healthcare system in most of the countries are overwhelmed due to the large number of patients with moderate-severe disease requiring hospitalization. The proportion of diagnosed asymptomatic infections reported are between 1 - 19·8%.^(21-25)^ The initial reports from China gave a frequency of 1% whereas, a much higher proportion at 19·8% was reported from the Republic of Korea.^(21,26)^ Asymptomatic ratio among evacuated Japanese nationals from China is estimated at 30·8%.^(24)^ This variation is due to the different strategies of surveillance and testing adopted in various settings. Our findings are similar to another reported study from India, although on a smaller sample size.^(14)^ Further, asymptomatic infections were more likely among patients with comorbidities with odds ratio of 2·66. This is consistent with another report of silent infections among young adults without comorbidities.^(25)^ The finding of 47% asymptomatic infections in the present study has a bearing on pandemic control strategies.

There is sparse literature on disease course and progression to severe disease among patients with mild disease and asymptomatic infection. In a study from Korea, out-of-hospital cohort treatment of COVID-19 patients with mild symptoms reported disease worsening in 2·3% patients. Patients with severe disease and underlying chronic severe medical conditions were excluded.^(27)^ In our study, none of the patients had disease progression to moderate-severe disease. Up triaging was required in 1·7% of the patients due to worsening of the underlying comorbidities. This data is reassuring that COVID-19 patients with mild disease or asymptomatic infections can be managed outside hospital settings. This will help in optimum utilization of healthcare facilities in resource-limited settings facing COVID-19 pandemic. This will also free up precious hospital beds for management of patients with moderate to severe disease.

This study has several limitations. First, there is sparse literature on mild cases and our patients were younger compared to other reported studies, so direct comparison of clinical manifestations and disease progression is to be done cautiously. However, our patients had a significant burden of comorbidities for their age, which is an important risk factor for severe disease. Secondly, this being a retrospective study there are inherent limitations. Daily symptom screen data was lacking in many cases leading to their exclusion, laboratory data was incomplete particularly with respect to markers like serum ferritin levels and C-reactive protein. Thirdly, there was a selection bias owing to aggressive contact tracing and admission of close contacts for testing. This resulted in a large number of asymptomatic patients being recruited for the study. Nevertheless, to the best of our knowledge, this is the largest report of clinical manifestations and disease course among admitted patients with mild COVID-19 and the data will be useful for policy makers.

## Conclusion

Patients with mild disease at presentation have a stable disease course and can be managed outside the hospital setting. Adequate care must be taken for comorbid conditions. A large proportion of patients are asymptomatic throughout the course of infection and patients with comorbidities are more likely to be symptomatic.

## Data Availability

Data collected and include in the manuscript is with chief investigators(RK and MS). It will be shared online if required.

